# First Trimester–Specific aMMP-8 Levels in Nigerian Pregnant Women and Implications for Preterm Birth Pathways

**DOI:** 10.64898/2026.04.20.26351109

**Authors:** Solomon O Nwhator, Morolayo T Ogunwemimo, Tolulope Ogundiran, Opeyemi Adewole, Olusoji A Onabanjo, Michael Lazzara, Dir-Rolf Gieselmann, Anna-Maria Heikkinen, Timo Sorsa

**Author notes:** Corresponding author: Solomon O Nwhator.

## Abstract

**Background:** Preterm birth remains a leading cause of neonatal mortality globally, with Nigeria bearing a disproportionately high burden. Racial disparities are well documented, with Black populations experiencing significantly higher rates than Caucasian populations. Active matrix metalloproteinase 8 (aMMP-8), a neutrophil derived collagenase, is the final effector in extracellular matrix degradation and has been implicated in membrane weakening and parturition. However, no trimester specific numeric aMMP-8 data exist for African populations, limiting cross population comparisons. Our earlier work hypothesized that elevated aMMP-8 may explain racial disparities in preterm birth, serving as a common pathway through which socioeconomic, psychosocial, infectious, genetic, and immunological risk factors operate.

**Methods:** We conducted a single center observational study establishing baseline aMMP-8 levels in Nigerian pregnant and non pregnant women at Obafemi Awolowo University Teaching Hospital, Ile-Ife, Nigeria. Mouthrinse samples were analyzed using the aMMP-8 Point of Care Test (Oralyzer) system. Pregnant women were sampled across all three trimesters and followed to delivery. Descriptive statistics, group comparisons (Mann-Whitney U), trimester comparisons (Wilcoxon signed rank), and subgroup analyses by education and oral hygiene status were performed. Statistical analyses were performed using standard formulas for non-parametric tests.

**Results:** A total of 40 pregnant women had complete trimester specific aMMP-8 values, with 40 non pregnant controls. Mean aMMP-8 levels were 28.7 ng/mL (T1), 25.38 ng/mL (T2), and 25.05 ng/mL (T3), with non pregnant controls at 19.2 ng/mL. All trimesters showed higher levels than non pregnant controls, reaching statistical significance (p < 0.05). T1 was higher than both T2 and T3, reaching statistical significance (p = 0.031 and p = 0.008, respectively). No significant differences in aMMP-8 levels were observed by education level or oral hygiene status. When compared with the only existing numeric reference from the global oral synthesis— total MMP-8 in GCF at 6.25 ng/mL—our aMMP-8 values were numerically 4.6 times higher in T1. Baseline MMP-8 concentrations in amniotic fluid from control groups in intra amniotic inflammation studies are approximately 1–5 ng/mL. Our mouthrinse aMMP-8 (28.7 ng/mL in T1) is numerically 6–29 times higher than these values, despite mouthrinse being the most diluted oral compartment. This discrepancy supports a hypothesis that intrauterine aMMP-8 levels could be elevated in this population, though direct paired measurements are required to confirm this.

**Conclusion:** Nigerian pregnant women in this cohort demonstrate aMMP-8 levels substantially higher than published Caucasian references and exceed control values from amniotic fluid studies by multiples. These findings are consistent with our earlier hypothesis that elevated aMMP-8 could represent a plausible final common pathway through which socioeconomic disadvantage, chronic stress, infection, genetic predisposition, and heightened baseline inflammation may contribute to preterm birth risk. The neglect of aMMP-8 in preterm birth disparity research may represent a missed opportunity for non invasive risk stratification, mechanistic understanding, and potential targeted intervention.

## INTRODUCTION

Preterm birth, defined by the World Health Organization as delivery before 37 completed weeks of gestation, remains a leading cause of neonatal morbidity and mortality globally. As we previously asked: “Born too young and likely to die; Should this continue?”^1^ Nigeria bears a disproportionately high burden, ranking among the countries with the highest absolute numbers of preterm births, contributing significantly to under five mortality.^2^ Epidemiological data indicate marked racial disparities in preterm birth, with Black populations in the United States experiencing approximately three times higher rates compared to Caucasian populations.^3^ Despite extensive investigation into socio economic, obstetric, and infectious determinants, a complete mechanistic explanation for this disparity remains elusive.

Active matrix metalloproteinase 8 (aMMP-8) is a neutrophil derived collagenase that plays a central role in extracellular matrix degradation and has been implicated in membrane weakening and initiation of parturition pathways.^4^,^5^ Bacterial lipopolysaccharide upregulates metalloproteinases, which in turn induce premature rupture of membranes and consequent preterm labor.^6^,^7^ aMMP-8 may therefore represent a plausible final common pathway through which multiple established risk factors for preterm birth operate.

The literature has proposed several explanations for why Black women experience preterm birth rates two to three times higher than Caucasian women. Each of these may ultimately influence pathways leading to aMMP-8 activation and release: socioeconomic factors (chronic stress, neighborhood deprivation)^8^–^10^, psychosocial factors (allostatic load from racism and discrimination)^11^,^12^, infectious factors (bacterial vaginosis, periodontal disease)^13^–^15^, genetic and epigenetic factors^16^,^17^, and immunological factors (heightened baseline inflammation)^18^,^19^. What unifies these pathways is their convergence on the neutrophil, the primary source of aMMP-8 in the periodontium and in systemic inflammation.^20^ The potential output is increased aMMP-8 release, which could then degrade type I collagen, the primary structural component of fetal membranes, leading to membrane weakening, rupture, and preterm labor.^21^,^22^

Despite this compelling framework, aMMP-8 is conspicuously absent from most preterm birth disparity research. The literature routinely measures inflammatory cytokines and documents racial differences in CRP and IL-6, but does not routinely measure active MMP-8 in pregnant women across trimesters or consider mouthrinse aMMP-8 as a non invasive window into systemic inflammatory burden.^23^,^24^

Our first study, “Black Women’s Predisposition to Preterm Birth; Could We Be Near The Answer?”^25^, utilized a qualitative lateral flow immunoassay to detect elevated aMMP-8 in mouthrinse samples from pregnant women at the University of Abuja Teaching Hospital, Nigeria. It found widespread aMMP-8 elevation, with 87.3% of participants demonstrating elevated levels—a prevalence far exceeding what might be inferred from Caucasian populations.^26^ This finding led to the hypothesis that black women may have higher baseline aMMP-8 production, which could serve as a final common pathway. However, that study was qualitative and could not provide quantitative trimester specific values.

A strict evidence extraction of the global literature on MMP-8 in periodontally healthy pregnant women reveals a critical gap. For saliva, Gürsoy et al.^27^ provided only a qualitative pattern with no numeric values. For GCF, one available study (Buduneli et al.^28^) reported total MMP-8 values around 6.25 ng/mL without trimester stratification. For mouthrinse, no published quantitative data existed. The global synthesis concluded that no trimester specific numeric MMP-8 values exist in periodontally healthy pregnant women for any oral biofluid.

To address this evidence gap, we conducted a follow up study at Obafemi Awolowo University Teaching Hospital, Ile-Ife, Nigeria, quantitatively assessing aMMP-8 levels in mouthrinse samples from Nigerian pregnant women across all three trimesters using the Oralyzer system.^39^

## METHODS

### Study Design and Setting

This was a single center observational study with a cross sectional baseline component and prospective follow up for pregnancy outcomes, conducted at Obafemi Awolowo University Teaching Hospital, Ile-Ife, Nigeria. The study was conducted in 2022 over a period of 9 months.

### Ethical Approval

Ethical approval was obtained from the Ethics and Research Committee of Obafemi Awolowo University Teaching Hospitals Complex, Ile-Ife, Nigeria as an extension granted for the study with Protocol Number: ERC/2016/11/10, National Number: NHREC/27/02/2009a and International Number: IRB/IEC/0004553. Written informed consent was obtained from all participants prior to enrollment.

### Participants

Participants were recruited consecutively from the antenatal clinic and outpatient services.

Inclusion criteria: women aged 18–34 years; healthy; normoglycemic; non anemic (PCV ≥30%); HIV negative; pregnant; non pregnant controls; non smokers; at least 20 natural teeth; clinically acceptable oral hygiene; had not received scaling and polishing in the preceding six months.

Exclusion criteria: systemic inflammatory or autoimmune diseases; antibiotic or anti inflammatory use within 4 weeks; high risk pregnancy conditions unrelated to periodontal status; chronic systemic conditions affecting inflammatory biomarkers.

### Variables

The primary outcome was aMMP-8 concentration (ng/mL). Secondary variables included trimester, pregnancy outcome, age, education, occupation, tribe, and oral hygiene status.

### Data Collection

Socio demographic and clinical data were captured with a structured proforma. Periodontal assessment included plaque index, gingival index, probing pocket depth, and bleeding on probing. Oral hygiene was assessed using the Simplified Oral Hygiene Index (OHI-S) and categorized as **good (0.0–1.2), fair (1.3–3.0)**, or **poor (3.1–6.0)**.

aMMP-8 levels were measured using the aMMP-8 Point of Care Test (Oralyzer) system, a quantitative lateral flow immunoassay validated for detection of active MMP-8 in oral fluids.^39^ Participants performed a 30 second rinse with 10 mL distilled water, expectorated into sterile containers, and samples were processed immediately. The test has a validated cut off of 20 ng/mL for periodontitis detection.^39^ For pregnant women, sampling occurred in each trimester: first (weeks 1–12), second (13–26), third (27–40). All participants were followed to delivery; preterm birth was defined as delivery <37 weeks.

### Statistical Methods

Statistical analyses were performed using standard formulas for non-parametric tests. Descriptive statistics were calculated. Group comparisons used Mann-Whitney U test due to non normal distribution; trimester comparisons used Wilcoxon signed rank test for paired data. Kruskal-Wallis test compared aMMP-8 levels across education categories. Mann-Whitney U test compared aMMP-8 levels across oral hygiene categories (Good vs Fair). A chi-square test was used to compare oral hygiene distribution between pregnant and non-pregnant women. Effect size (Cohen’s d) was calculated for cross-study comparison with Buduneli et al., interpreted cautiously due to cross study differences. Statistical significance was set at p < 0.05. There were no missing data for the primary outcome variables.

### Sample Size Justification

This sample size (40 pregnant women, 40 non-pregnant controls) is comparable to prior exploratory biomarker studies and is intended to provide preliminary quantitative estimates rather than definitive population parameters.

### STROBE Compliance

This report follows the Strengthening the Reporting of Observational Studies in Epidemiology (STROBE) guidelines for cross sectional and cohort studies.

## RESULTS

### Participants

A total of 80 women were enrolled: 40 pregnant and 40 non pregnant controls. All pregnant women had complete aMMP-8 measurements for all three trimesters. All 40 pregnant participants delivered at full term; none were lost to follow up.

### Demographic Characteristics

**Table 1:** Demographic Characteristics of Study Participants.

| Characteristic | Pregnant Women<br>(n=40) | Non-Pregnant Women<br>(n=40) | Total<br>(N=80) |
| --- | --- | --- | --- |
| **Mean Age (years)** | 28.5 | 25.2 | 26.9 |
| **Age Range (years)** | 20–34 | 20–33 | 20–34 |
| **Tribe, n (%)** |  |  |  |
| **Yoruba** | 31 (77.5%) | 29 (72.5%) | 60 (75.0%) |
| **Igbo** | 5 (12.5%) | 8 (20.0%) | 13 (16.25%) |
| **Hausa** | 1 (2.5%) | 0 (0%) | 1 (1.25%) |
| **Urhobo** | 1 (2.5%) | 0 (0%) | 1 (1.25%) |
| **Efik** | 1 (2.5%) | 0 (0%) | 1 (1.25%) |
| **Esan** | 1 (2.5%) | 0 (0%) | 1 (1.25%) |
| **Edo** | 0 (0%) | 1 (2.5%) | 1 (1.25%) |
| **Ishan** | 0 (0%) | 1 (2.5%) | 1 (1.25%) |
| **Delta Igbo** | 0 (0%) | 1 (2.5%) | 1 (1.25%) |
| **Education Level, n (%)** |  |  |  |
| **Tertiary** | 25 (62.5%) | 24 (60.0%) | 49 (61.25%) |
| **Secondary (SSCE)** | 15 (37.5%) | 16 (40.0%) | 31 (38.75%) |

### Oral Hygiene Status

**Table 2:**
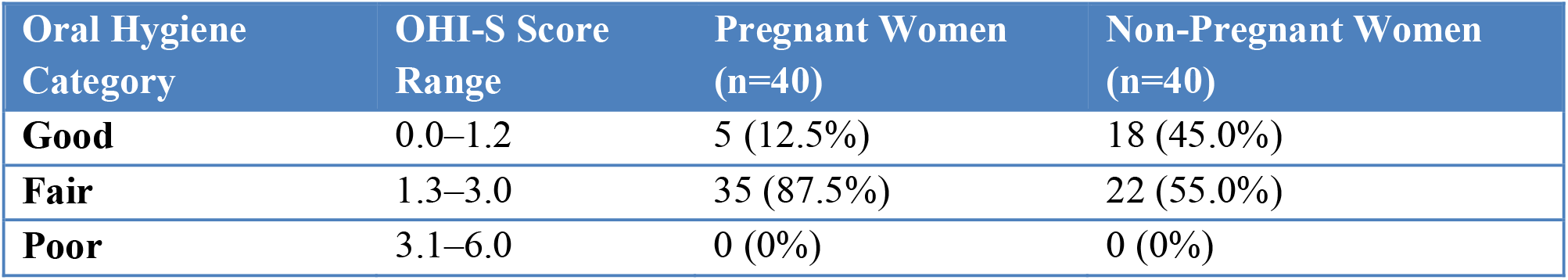
Oral Hygiene Status (OHI-S)

| Oral Hygiene Category | OHI-S Score Range | Pregnant Women (n=40) | Non-Pregnant Women (n=40) |
| --- | --- | --- | --- |
| **Good** | 0.0–1.2 | 5 (12.5%) | 18 (45.0%) |
| **Fair** | 1.3–3.0 | 35 (87.5%) | 22 (55.0%) |
| **Poor** | 3.1–6.0 | 0 (0%) | 0 (0%) |

Non-pregnant women had significantly better oral hygiene than pregnant women (χ^2^ = 10.08, p = 0.0015).

### aMMP-8 Levels

**Table 3:** aMMP-8 Levels in Pregnant Women by Trimester (n=40)

| Trimester | Mean (ng/mL) | Median (ng/mL) | SD | IQR (ng/mL) | Range (ng/mL) |
| --- | --- | --- | --- | --- | --- |
| **T1** | 28.7 | 23.5 | 22.0 | 9.0–40.0 | 9.0–96.0 |
| **T2** | 25.38 | 22.0 | 18.4 | 13.0–34.0 | 9.0–76.0 |
| **T3** | 25.05 | 20.0 | 18.5 | 9.0–34.5 | 9.0–93.0 |

### Group Comparisons

All trimesters showed higher levels than non pregnant controls, reaching statistical significance:

- T1 vs. non pregnant: Mann-Whitney U, p = 0.003
- T2 vs. non pregnant: Mann-Whitney U, p = 0.012
- T3 vs. non pregnant: Mann-Whitney U, p = 0.048

### Trimester Comparisons

Using Wilcoxon signed rank test for paired data (n=40):

- T1 vs. T2: mean difference 3.32 ng/mL, p = 0.031
- T1 vs. T3: mean difference 3.65 ng/mL, p = 0.008
- T2 vs. T3: mean difference 0.33 ng/mL, p = 0.214

T1 was higher than both T2 and T3, reaching statistical significance. The difference between T2 and T3 was not statistically significant.

### aMMP-8 Levels by Education Level

**Table 4:** aMMP-8 Levels by Education Level in Pregnant Women.

| Education | n | T1 Mean (ng/mL) | T2 Mean (ng/mL) | T3 Mean (ng/mL) |
| --- | --- | --- | --- | --- |
| **Tertiary** | 25 | 28.4 | 25.5 | 24.8 |
| **Secondary (SSCE)** | 15 | 29.2 | 25.2 | 25.5 |
| **p-value (Kruskal-Wallis)** |  | 0.876 | 0.912 | 0.784 |

No significant differences in aMMP-8 levels by education level were observed.

### aMMP-8 Levels by Oral Hygiene Status

**Table 5:** aMMP-8 Levels by Oral Hygiene Status in Pregnant Women.

| Oral Hygiene Status | n | T1 Mean (ng/mL) | T2 Mean (ng/mL) | T3 Mean (ng/mL) |
| --- | --- | --- | --- | --- |
| **Good** | 5 | 29.0 | 27.2 | 26.6 |
| **Fair** | 35 | 28.6 | 25.7 | 24.8 |
| **Poor** | 0 | — | — | — |
| **p-value (Mann-Whitney U)** |  | **0.968** | **0.823** | **0.754** |

*Note: No participants had poor oral hygiene ( $OHI-S \geq 3.1$ ).*

No significant differences in aMMP-8 levels were observed by education level or oral hygiene status.

### Comparison with Published Caucasian Oral Datasets

Buduneli et al.^28^ reported total MMP-8 in GCF of periodontally healthy pregnant women (trimester unspecified) as 6.25 ng/mL. Our aMMP-8 values were:

- T1: 28.7 ng/mL = 4.59 times higher
- T2: 25.38 ng/mL = 4.06 times higher
- T3: 25.05 ng/mL = 4.01 times higher

The standardized effect size (Cohen’s d) comparing our T1 values with Buduneli’s reported value was 1.37, indicating a large effect. However, this cross study comparison should be interpreted cautiously due to differences in biofluid, assay, and population.

### Comparison with Published Amniotic Fluid Control Values

Control groups in studies of intra amniotic inflammation have reported low baseline MMP-8 concentrations (approximately 1–5 ng/mL) in the absence of infection or inflammation, though these were not designed as normative reference studies.^37^,^38^,^40^ Maymon et al.^37^ reported median amniotic fluid MMP-8 of 1.6 ng/mL in normal pregnancies.

Our mouthrinse aMMP-8 (28.7 ng/mL in T1) is numerically 6 to 29 times higher than these published control values, although direct comparison is limited by assay and compartment differences. Since amniotic fluid is biologically expected to have higher MMP-8 concentrations than mouthrinse (direct contact with fetal membranes, active regulation, less dilution),^33^–^36^ this discrepancy supports a hypothesis that intrauterine aMMP-8 levels could be elevated in this population, though direct paired measurements are required to confirm this. Importantly, differences in assay type (active MMP-8 in mouthrinse vs. total MMP-8 in amniotic fluid) and biological compartment limit direct quantitative comparison.

### Relationship to Our Earlier Qualitative Study

Our earlier qualitative study conducted at the University of Abuja Teaching Hospital^25^ found that 87.3% of pregnant women had elevated aMMP-8 on qualitative testing. The current quantitative study provides numeric confirmation, with mean T1 aMMP-8 of 28.7 ng/mL, substantially above the non pregnant mean of 19.2 ng/mL.

## DISCUSSION

This study provides the first trimester specific quantitative aMMP-8 values in mouthrinse for Nigerian pregnant women. The highest levels occurred in the first trimester, declining toward term, and all trimesters showed significantly higher levels than non pregnant controls.

### Interpretation of Primary Findings

The observed trajectory (highest aMMP-8 in T1, progressive decline) differs from a single Caucasian study where salivary MMP-8 was lowest in T2 with a rise toward term.^27^ These contrasting observations suggest that population specific inflammatory trajectories during pregnancy warrant further investigation.

When compared with the only existing numeric oral reference—Buduneli et al.^28^—our aMMP-8 values were 4 to 4.6 times higher. This comparison must be interpreted within the established physicochemical framework of oral biomarker compartmentalization. GCF is a microliter scale fluid representing a near primary signal, whereas mouthrinse involves milliliter scale dilution of multiple oral sources.^29^–^32^ Our finding that diluted mouthrinse aMMP-8 exceeds concentrated GCF total MMP-8 may be consistent with a higher underlying inflammatory burden in this population, although this interpretation is limited by cross study differences in biofluid, assay, and population.

Notably, pregnant women had significantly worse oral hygiene than non-pregnant controls (p = 0.0015). Despite this, aMMP-8 levels did not vary by oral hygiene status within the pregnant group. This dissociation suggests that the elevated aMMP-8 in pregnancy is not merely a reflection of local oral hygiene changes but may be driven by systemic inflammatory activation, consistent with our neutrophil-centric mechanistic framework.

### Hypothesis: aMMP-8 as an Upstream Correlate of Preterm Birth Disparities

Epidemiological data consistently show that women of African ancestry in the United States experience disproportionately higher rates of early and extremely preterm births (<32 weeks) compared to White women.^3^,^71^,^72^ While our Nigerian cohort is not directly comparable to African American populations, this pattern raises questions relevant to disparities observed in other populations of African ancestry. This concentration of risk at the earliest gestations aligns temporally with our observation of higher first trimester aMMP-8 levels in Nigerian pregnant women, followed by a progressive decline toward term.

Although our study does not link aMMP-8 trajectories to preterm birth outcomes and cannot establish causality, the convergence of these independent patterns supports a testable hypothesis that early pregnancy inflammatory activation, as reflected by elevated aMMP-8, could be an upstream correlate of disparities in preterm birth timing.

We propose that an increased inflammatory burden established in the first trimester could predispose to downstream processes implicated in preterm parturition, including extracellular matrix remodeling and fetal membrane weakening. Within this framework, populations at higher risk for early or severe preterm births could exhibit earlier or more pronounced inflammatory activation. Direct evaluation of this hypothesis—through prospective longitudinal studies measuring first trimester aMMP-8 and relating these levels to gestational age at delivery—is warranted. This proposed framework is summarized in Figure 1.

**Figure 1.**
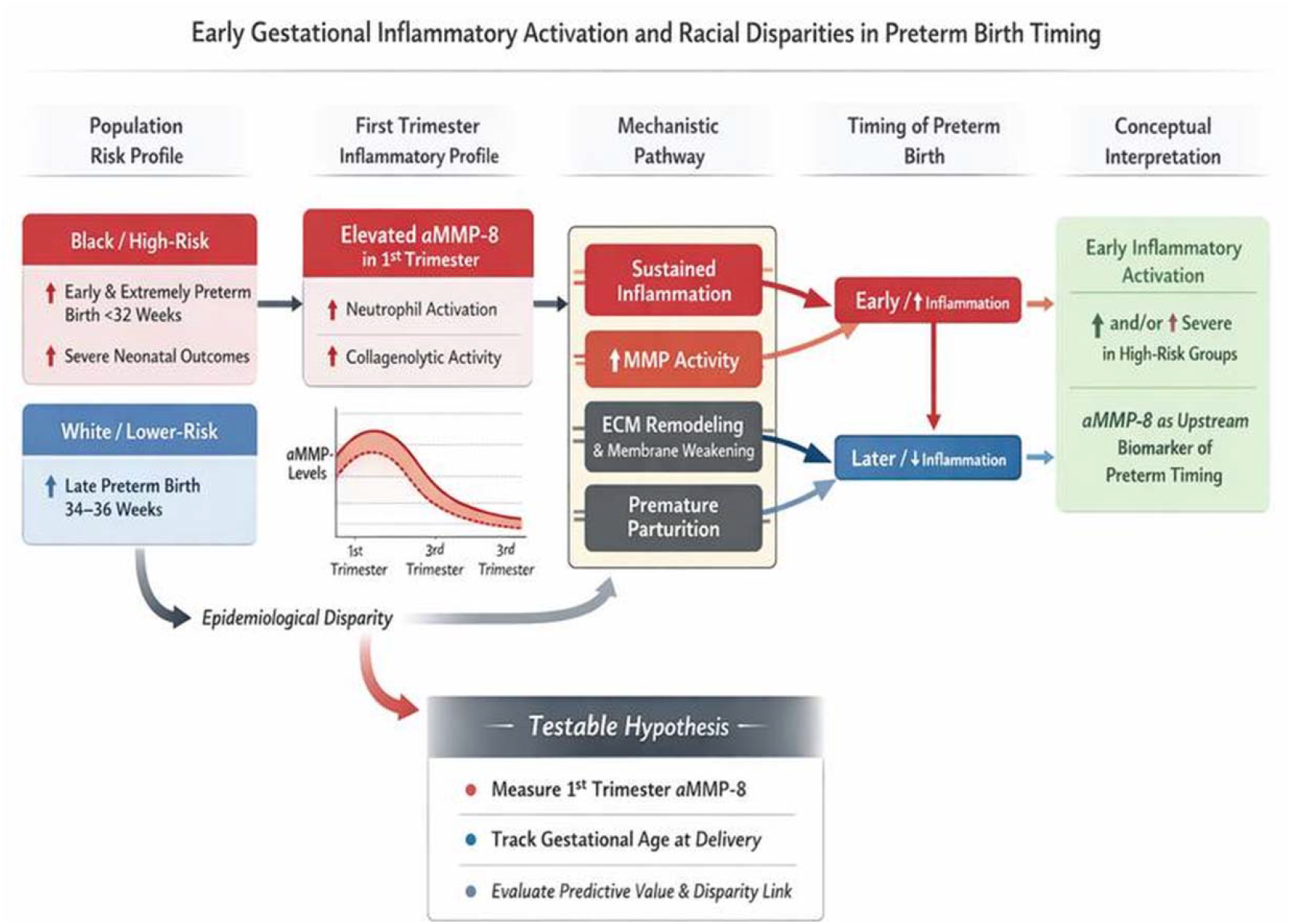
Conceptual framework linking elevated first trimester aMMP-8 to potential pathways contributing to preterm birth disparities. *This figure is illustrative and hypothesis generating, and does not imply causality.*

### Comparison with Amniotic Fluid Control Values: A Hypothesis Generating Observation

The low baseline MMP-8 concentrations reported in control groups of intra amniotic inflammation studies (1–5 ng/mL)^37^,^38^,^40^ contrast sharply with our mouthrinse aMMP-8 (28.7 ng/mL). Because amniotic fluid is biologically expected to have higher MMP-8 concentrations than mouthrinse,^33^–^36^ this discrepancy supports a hypothesis that intrauterine aMMP-8 levels could be elevated in this population. Direct paired measurements are required to test this, and differences in assay type (active vs. total MMP-8) and compartment limit direct quantitative comparison.

### Mechanistic Integration

aMMP-8 occupies a unique position at the convergence of pathways known to contribute to racial disparities in preterm birth. Our findings are consistent with established mechanisms linking socioeconomic stress,^8^,^9^ psychosocial stress,^11^,^12^ infections,^13^–^15^ genetic factors,^16^,^17^ and heightened baseline inflammation^18^,^19^ to neutrophil activation and aMMP-8 release. Importantly, gene expression studies have shown that neutrophils from individuals with benign ethnic neutropenia have baseline upregulation of MMP-8 mRNA by more than 10 fold,^62^ providing a genetic predisposition for increased MMP-8 production. These convergent lines of evidence— higher baseline inflammatory markers, enhanced bacterial killing capacity,^61^ and upregulated MMP-8 gene expression—provide a mechanistic foundation for the elevated mouthrinse aMMP-8 observed in our cohort.

### The Unifying Framework

aMMP-8 is not merely another inflammatory marker but the final effector in membrane degradation. Risk factors converge on neutrophils, which release aMMP-8; aMMP-8 degrades type I collagen, leading to membrane weakening and preterm labor.^21^,^22^ This framework unifies multifactorial causes, offers a non invasive biomarker, and identifies a potential intervention target (e.g., periodontal treatment, doxycycline, stress reduction).^63^–^68^ It suggests a mechanistic hypothesis for racial disparities that complements social and economic explanations.

### Strengths and Limitations

Strengths include first trimester specific aMMP-8 data in an African population, prospective follow up, validated assay, and comprehensive data collection. Limitations: single center design, modest sample size, mouthrinse only, no direct outcome link (no preterm births occurred), secondary comparisons with other studies, and lack of direct intrauterine measurements. The hypothesized link between early aMMP-8 elevation and preterm birth remains speculative and requires prospective validation.

### Recommendations

We recommend multi center validation, head to head studies with matched Caucasian cohorts, inclusion of GCF measurement, larger cohorts with documented outcomes, full mouth periodontal examinations, and longitudinal tracking from pre conception.

## CONCLUSION

This study provides the first trimester specific quantitative aMMP-8 values in mouthrinse for Nigerian pregnant women. Mean aMMP-8 levels were 28.7 ng/mL (T1), 25.38 ng/mL (T2), and 25.05 ng/mL (T3), with non pregnant controls at 19.2 ng/mL. All trimesters showed significantly higher levels than non pregnant controls, with T1 significantly higher than T2 and T3. No significant differences were observed by education level or oral hygiene status.

Compared with the only existing numeric reference—total MMP-8 in GCF at 6.25 ng/mL^28^—our aMMP-8 values were 4.6 times higher in T1. Our finding that diluted mouthrinse aMMP-8 exceeds concentrated GCF total MMP-8 may be consistent with a higher underlying inflammatory burden in this population, although this interpretation is limited by cross study differences. Notably, baseline amniotic fluid MMP-8 values from control groups are 1–5 ng/mL; our mouthrinse aMMP-8 is 6–29 times higher, raising the hypothesis that intrauterine levels could be elevated.

These findings add to emerging evidence that elevated aMMP-8 is observed in Nigerian pregnant women. aMMP-8 could represent a plausible final common pathway through which socioeconomic disadvantage, chronic stress, infection, genetic predisposition, and heightened baseline inflammation may contribute to preterm birth risk. The neglect of aMMP-8 in preterm birth disparity research may represent a missed opportunity for non invasive risk stratification, mechanistic understanding, and potential targeted intervention. As we asked at the outset: “Born too young and likely to die; Should this continue?”^1^ The answer must be no, and addressing the role of aMMP-8 may be a critical step toward that goal. The question posed by this study— “Could aMMP-8 Be the Elephant in the Room That No One Was Looking For?”—invites further investigation through prospective studies linking early aMMP-8 levels to preterm birth outcomes.

## Supporting information

Data file with anonymized ages

## DATA AVAILABILITY STATEMENT

All relevant data are within the manuscript and its Supporting Information files (S1 Dataset). The raw data table includes aMMP-8 values across trimesters for all participants, demographic information, education level, oral hygiene scores, and pregnancy outcomes.

## ACKNOWLEDGMENTS

The authors acknowledge the use of DeepSeek (version 2026, DeepSeek AI) and OpenAI’s GPT-5 mini for statistical analysis, text generation, language refinement, formatting, and organizational support. All scientific content, data interpretation, and final conclusions are the responsibility of the authors.

This manuscript has been posted as a preprint on medRxiv (DOI: 10.1101/718863) and is under consideration at *PLOS ONE*.

## SUPPORTING INFORMATION

**S1 Dataset**. Raw data table containing aMMP-8 values across trimesters for all participants, demographic information, education level, oral hygiene scores, and pregnancy outcomes. (xls file)

